# Comparing the predictive value of suicide risk screening to the detection of suicide risk using electronic health records in an urban pediatric emergency department

**DOI:** 10.1101/2020.12.27.20248829

**Authors:** E.E. Haroz, C. Kitchen, P.S. Nestadt, H.C. Wilcox, J.E. DeVylder, H. Kharrazi

**Affiliations:** Center for American Indian Health, Department of International Health, Johns Hopkins Bloomberg School of Public Health, Baltimore, MD; Center for Population Health IT, Department of Health Policy and Management, Johns Hopkins Bloomberg School of Public Health, Baltimore, MD; Department of Psychiatry and Behavioral Sciences, Johns Hopkins School of Medicine, Baltimore, MD; Department of Mental Health, Johns Hopkins Bloomberg School of Public Health, Baltimore, MD; Graduate School of Social Service, Fordham University, New York, NY; Division of Health Sciences Informatics, Johns Hopkins School of Medicine, Baltimore, MD

**Keywords:** Suicide Screening, Suicide Prevention, Machine Learning

## Abstract

**Objective:** To compare the accuracy, sensitivity and utility of brief screening to predictive modeling for identifying suicide-related outcomes in a pediatric emergency department. Our hypothesis was that predictive modeling would be more accurate and useful compared to brief screening.

**Methods:** This was a retrospective cohort study at an urban pediatric Emergency Department (PED) in the United States. Patients were aged 8 to 18 years old who presented from January 1, 2017 to June 30, 2019. Predictors included positive suicide risk screen and/or demographic, medical and mental health diagnostic Electronic Health Record (EHR) data at the time of visit. The main outcome was subsequent PED visit with suicide-related presenting problem (i.e. ideation or attempt) within a 3-month follow-up period.

**Results:** Of the *N=*13420 individuals, *n=*141 had a subsequent suicide-related ED visit. Approximately 63% identified as Black Non-Hispanic. Results showed that a model based only on EHR data performed only slightly worse than the ASQ alone. Combining ASQ screening and EHR data resulted in a 17.4% improvement in sensitivity and 13.4% increase in AUC compared to the ASQ alone. The LASSO models indicated a diagnosis of major depressive disorder in addition to the ASQ improved the AUC by 9%.

**Conclusions, Implications, Future Directions:** Our findings show that EHR data is helpful either in the absence or as an addition to brief suicide screening. To our knowledge, this is the first study to compare brief screening to EHR based predictive modeling and adds to our understanding of how best to identify youth at risk of suicidal behaviors in clinical care settings.

## Introduction

Since 2014, suicide has been the 2^nd^ leading cause of death among youth aged 10-14 and 15-24-years old in the United States.^1^ Suicide rates have increased in nearly every state since 1999, with female and Black youth showing especially marked increases over this time period.^2–5^ Moreover, 17.2% of high school students nationwide have reported seriously considering attempting suicide in the last year, and 7.4% of students report having attempted suicide in the past 12 months.^6^ Identifying youth most at risk of attempting or dying by suicide is a key part of any preventative approach. The timely identification of those at risk and provision of appropriate and effective care could have a major impact on driving down suicide rates.

Healthcare institutions serve as a critical setting for suicide risk detection. First, healthcare settings area potential point of contact for someone at risk of attempting or dying by suicide. Several studies focused on examining medical records in the year prior to among suicide decedents have found high rates of utilization among those that died.^7,8^ For example, Ahmedani et al.,^7^ showed that in the month prior to their death, half of all suicide decedents received healthcare services at least once. Second, the acute time period following discharge from a healthcare facility is often characterized by elevated risk for suicide attempt and death.^9–11^ Chung et al.^10^ showed that across 100 studies, the rate of suicide during the first 3 months after discharge from a psychiatric facility was approximately 100 times higher than the global suicide rate. The increase in risk following healthcare contact is also seen among patients discharged from non-psychiatric settings, including emergency departments (EDs), as well.^9^

The identification of suicide risk at healthcare facilities is a major priority in suicide prevention efforts. However, in recent years two distinct approaches have emerged regarding how best to do this. The first approach emphasizes the use of short, validated screening measures. The Joint Commission (TJC) requires screening for suicide risk among all patients being evaluated or treated for behavioral health conditions with a validated screening tool, to be in compliance with the National Patient Safety Goal 15.01.01.^12^ More broadly, the US Action Alliance’s “Zero Suicide” (ZS) model for suicide prevention in healthcare systems includes universal screening for suicide risk at all points of care in a healthcare setting as one of several essential elements.^13^ Studies have shown that use of brief screeners is feasible,^14^ that positive screens are significantly associated with subsequent suicide-related hospital visits,^15^ and that screening may double detection of those most at risk.^15,16^ However, implementation of screening involves several important hurdles, including generating buy-in from hospital administrators, programming of screeners into the electronic health record (EHR), training of staff in screening procedures and appropriate triage actions, and ongoing monitoring of fidelity to the instrument and process.^14,17–19^ Many screeners, which generally center on the question of suicidal ideation, can suffer from both high rates of false positives as well as even more dangerous false negatives, which can create a false sense that risk has been appropriately ruled in or out.^20^

Simultaneously, the field of suicide prevention has also been pursuing predictive modeling as a method for identifying individuals at risk for attempting or dying by suicide. The availability of large health care datasets such as the EHR,^21^ combined with advances in machine learning and an increased understanding of the complexities of co-occurring risk factors for suicide,^22^ has led to a growing number of studies aimed at developing, validating and deploying prediction models as routine care to identify individuals at risk for subsequent attempts and deaths.^23–27^ Accuracy of these models is generally high;^23^ however, these models have rarely been implemented as routine care in health systems^28^ as model deployment is often complicated by factors related to preparing data, developing clinician friendly tools and notifications, and ethical and regulatory issues.^29,30^

The parallel development and enthusiasm for both universal screening and predictive modeling begs several questions: First, are either of these approaches more accurate at identifying those at risk? Second, how often do these methods agree? Third, how can these methods work in tandem; And, forth, what are the implications for implementation related to these two approaches? The present study compared these two approaches to predicting subsequent suicide related outcomes among a sample of patients aged 8-18 years who presented to the Johns Hopkins Hospital (JHH) pediatric emergency department (PED) from January 1, 2017 to June 30, 2019. The aims were to (1) compare a brief screening measure (the Ask Suicide-Screening Questions; ASQ^31^) to an EHR-based predictive modeling approach on accuracy, sensitivity and utility for identifying those at risk of subsequent return to the PED for suicide-related reasons; and (2) measuring the added value of integrating EHR with ASQ for identify subsequent PED suicide-related visits among our sample. Our hypotheses were that EHR-based predictive modeling would provide similar accuracy, sensitivity and utility as brief suicide screening; and the combination of risk screening and EHR-based predictive modeling would perform the best. To our knowledge, no study has directly compared EHR-based predictive modeling methods to brief suicide risk screeners in terms of their ability to identify those at risk for future suicidal behaviors. Findings from this study will inform our understanding of how best to identify individuals at risk of suicide, as well as implications for implementing such approaches in clinical care settings.

## Methods

The study was a retrospective cohort study of all patients seen at Johns Hopkins Children’s Center’s PED from January 1, 2017 to June 30, 2019. Johns Hopkins Children’s Center’s PED is part of an urban academic medical center located in Baltimore, MD and provides over 30,000 ED visits a year. The medical record review and analysis included in this study was approved by the Johns Hopkins School of Medicine Institutional Review Board (IRB). Patients did not need to provide informed consent for screening as all data were collected during routine care. All reporting for this study followed the Strengthening the Reporting of Observational Studies in Epidemiology (STROBE) guidelines for cohort studies.

### Screening Assessment

Universal screening for risk of suicide was accomplished using the Ask Suicide-Screening Questions (ASQ) of all patients ages 10 and older and patients ages 8 and 9 presenting with a behavioral or psychiatric chief complaint. The ASQ is a freely available, brief, validated tool designed for use with patients ages ten years or above.^31^ It includes four screening questions that take 20 seconds to administer.^31^ A positive screen is indicated if a positive (i.e. “yes”) response is provided to any of the four questions. If the patient screens positive, a fifth question is asked aimed at understanding acuity of the risk. The ASQ was administered by nurses in the PED during triage. If a patient screened positive, the protocol required that th ED physician was notified and the patient was provided with additional evaluations and referrals as appropriate, although how often this actually happened was variable.^32^

### Measures

Our outcomes of interest are defined as a positive instance of suicide ideation or attempt at PED visit within three and twelve months of the index visit, independent of responses obtained as part of ASQ screening. We did not rely only on International Classification of Diseases (ICD) codes (e.g. E950-E958) as these were seldom found for the same ED encounters (90 of 25,067 ED encounters between 1/1/2017 and 6/30/2019). Thus, we defined instances of PED suicide ideation or attempt on recorded principal diagnosis description and chief complaint as documented in the EHR, for those missing ICD codes. These fields contained free-text values that reflect reasons for the patient being seen in the ED, and tended to be very brief (on average 13 to 32 characters, or 39-100 max). Our outcome was a binary indicator for pattern matched terms “suicide”, “suicidal”, “suicide ideation”, “suicide gesture”, “self-cutting”, “self-harm”, “self-injury”, “self-injurious”, “self-inflicted”, “self-mutilation”, “self-mutilating”, “suicide attempt”, and “attempted suicide” on either PED chief complaint or presenting diagnosis. As a sensitivity analysis, we compared our pattern matched outcome measure to manually scored suicidal behaviors for overlapping patients in data from DeVylder et al.^15^

Predictors of interest focused on both acute and chronic psychiatric and medical comorbidities diagnosed at the time of PED visit. Twenty-five medical and psychiatric condition categories were obtained from the Centers for Medicare and Medicaid Services (CMS) Chronic Conditions Warehouse (CCW) and were matched to ICD codes extracted from index PED visits. The CCW markers were chosen on the basis of prior utility in PED suicide risk prediction,^15^ plausibility for incidence within the pediatric population (i.e. excluding conditions such as stroke, hip fracture, osteoporosis), and potential relevance for other clinical settings. An additional 17 conditions and 4 comorbidity risk scores (i.e. Charlson comorbidity index) were obtained using the ‘comorbidity’ package^33^ for the R programming language (3.6.2). Additional markers were created for patient age, gender and whether a patient screened positive on the ASQ. We did not include an indicator for race and/or ethnicity for concern about propagating potential “race-based medicine.”^34^ The ASQ item 4 was also retained for use in a contrast model and consists of asking patients whether they have ever tried to kill themselves. In this manuscript, demographic and diagnostic information, including the Charlson weighted index of comorbidity, are referred to as “EHR” data. We then compared the value of selected EHR data to an ASQ positive screen or affirmative response on ASQ item 4 for predicting suicide-related ED visits.

### Data processing

Data preparation consisted of fitting available PED encounter level data to all patients seen in the PED between 1/1/2017 and 6/30/2019. The aggregation of encounter data required identifying case and control groups, defined by whether a patient had an ED visit for suicide ideation or attempts within 3 months of an initialized PED visit. To identify as many cases as possible, we first found all patients that had PED visits in the preceding three months of an ED visit with ideation or attempt. If a patient had multiple spans in which this condition could be met, only the first instances were retained. Relevant encounter data were aggregated within the observation interval such that the presence of any diagnostic condition was flagged positive, leading up to the visit containing ideation or attempt. ‘Controls’ were similarly identified and processed, with indexing on the first observed encounter date and included all single event PED visits. In this way, it was possible for ‘controls’ to have an initial instance of ideation or attempt, but *not* a subsequent one within three months of the indexed date. Finally, ‘control’ patients who had less than 3 months of observation (i.e. first visit after March, 30^th^ 2019) were removed from the analysis.

### Missing Data

Only missing values for the ASQ item 4 were necessary for modeling. These missing values were imputed using the ‘mice’ package in R.^35^ A total 663 values were imputed for ASQ item 4 using this method. For the other ASQ items missing values existed with progressively more missing values later in the questionnaire (ASQ 1: 102; ASQ 2: 630; ASQ 3: 620; ASQ 4: 663), suggesting that omitted values resulted from questions not being asked at the time of screening.

### Statistical Analysis

#### Exploratory analyses

We compared cases and controls on the basis of age, gender, race/ethnicity, ASQ screening outcome and subsequent ED visit outcome (i.e. ideation or attempt-related or not). All predictors and ASQ questions responses were loaded into point biserial correlation matrices, which aided in removal of collinear or empty variables. Our outcome sensitivity analysis found excellent overall accuracy (97.9%) when compared to previously manually coded outcomes in a similar dataset, but different study population (e.g. DeVylder et al.^15^). The pattern matched definition was also more inclusive, with 309 additional instances of suicidal ideation or attempt over those detected manually and only 6 instances where manual review identified suicide ideation or attempt where our definition did not.

#### Modeling

The three-month and 12-month analyses consisted of the construction of four models predicting ED visits for suicide ideation or attempt. These included: (1) ASQ positive screen, (2) ASQ positive screens and EHR content, (3) EHR content without the ASQ positive screen, and\ (4) The EHR content with item 4 of the ASQ. Model 4 was added, given the importance of a history of suicide attempt for future suicide risk^20^ and the relative ease of asking one question compared to five. Three-month analyses are presented in this paper, while twelve month results are included in the Appendix (see Appendix A1).

For all models, five-fold cross validation was used with binary logistic regression and LASSO regression. Binary logistic regressions were performed using the “cv” method with the train function of R’s caret library.^36^ LASSO was performed using cv.glmnet with a binomial link function and lambda set at one standard error above average during cross validation. Minimum classification thresholds were set to the probability of a randomly chosen patient having subsequent PED visit for ideation or attempt within three months (*p =* .01). Performance metrics were returned for model accuracy, sensitivity, specificity, positive predictive value, false positive rate and AUC (area under the curve). Model selection demanded consideration of sensitivity, parsimony and fit (AUC) given the high cost of false negative prediction. Variable importance and ROC (receiver operating characteristic) plots were generated to further interpret the performance of the binary logistic regression models.

### Secondary Analysis

The above definition of our outcome of interest in PED visits allows for the inclusion of an additional variable: whether a patient had a ED visit for ideation or attempt within the past three months (twelve months included in Appendix A1) for of their outcome. This was defined as the above pattern matched terms for presenting diagnosis description and chief complaint within the interval leading up to the outcome event. Binary logistic regression and LASSO models were developed with all predictors plus an indicator for ideation or attempt at index visit to inspect the added value of this one marker on model performance (See Appendix A2).

## Results

### Sample Characteristics

A total of 13 420 unique patients in the targeted age range were seen in the PED between January 1, 2017 and June 30, 2019 (**Table 1**). Among our sample, 7040 (52.5%) were female, 6280 (47.5%) were male, and the average age was 14.3 years old. The majority of the sample identified as Black Non-Hispanic (8438; 63%), while a quarter self-identified as White Non-Hispanic (3370; 25%), 914 (6.8%) as Hispanic, and 703 (5.2%) as other or more than one race. All patients seen in the PED were screened with the ASQ. Compliance with each individual ASQ question was high: 99.2% for question 1, 95.3% for question 2; 95.4% for question 3, and 95.1% for question 4. A total of 865 patients (6.4%) presented with an initial complaint of either suicidal ideation and/or suicide attempt during the timeframe for this analysis, from whom 141 patients had a subsequent suicide-related ED visit.

**Table 1.**
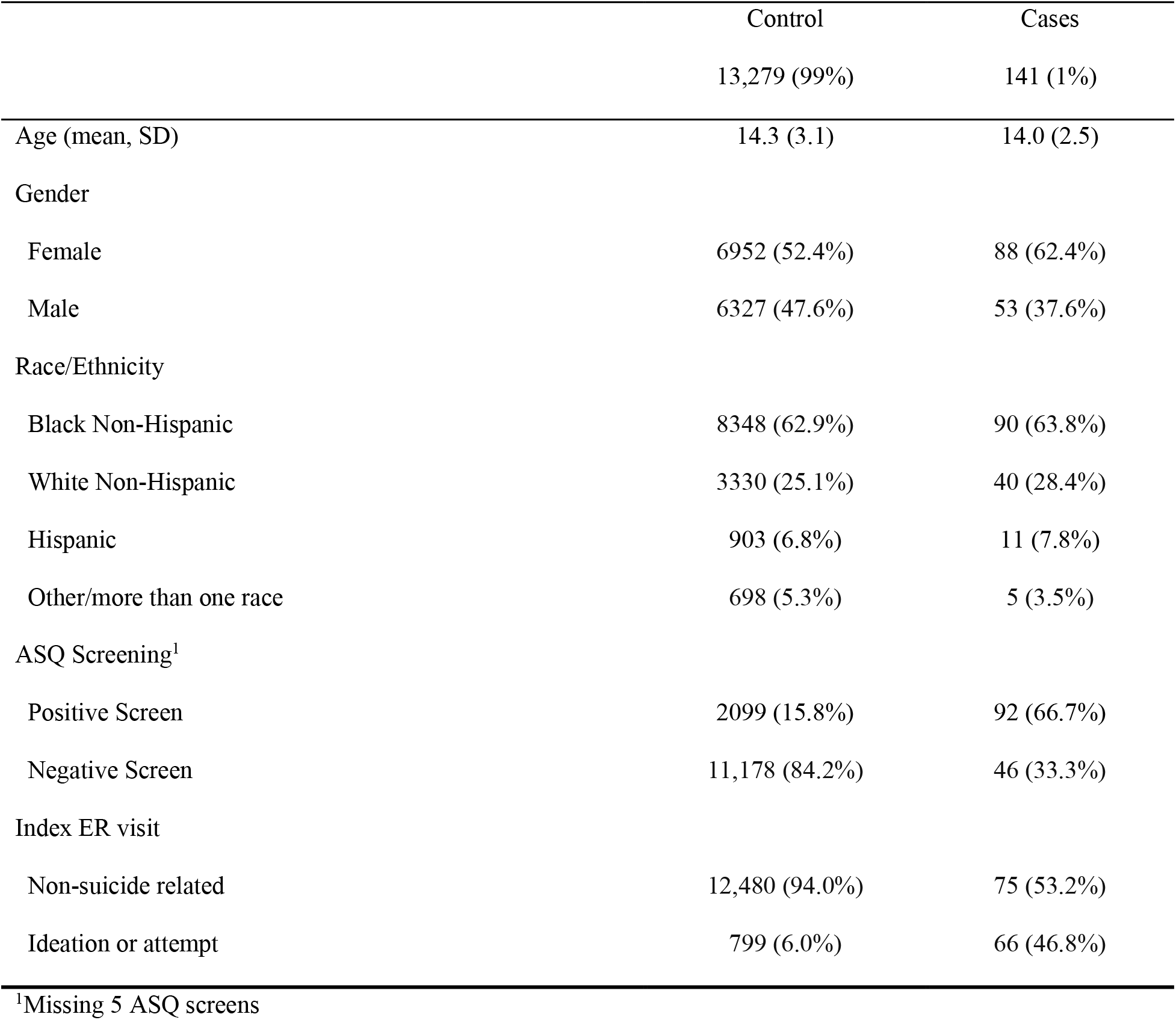
Demographic and clinical descriptive data by case status (*N =* 13,420)

### Prediction Models

**Table 2.** shows the comparison of binary logistic regression models based on accuracy, sensitivity, specificity, positive predictive value (PPV), false positive rate and AUC. Model 1, which includes just the binary predictor of ASQ positive vs. negative screen on subsequent suicide-related ED visit had the highest accuracy metric (0.840), but relatively low sensitivity (0.667) and the lowest value for AUC (0.754; **Figure 1**.). A model based only on data from the EHR (Model 3), performed only slightly worse than the ASQ alone as it had the lowest sensitivity (0.601) but slightly higher AUC value (0.775; **Figure 1**.). The best performing model was the combination of screening with the ASQ and use of EHR (Model 2). This model showed an accuracy of 0.825, sensitivity of 0.783 and AUC of 0.855 (**Figure 1**.). When EHR is added to the ASQ screening process, the sensitivity is improved by 17.4%, AUC by 13.4%, while there is a slight loss of specificity (−1.9%). Across all models negative predictive value (NPV) was higher than 0.99. False positive rates ranged from 15.8% for Model 1 to 17.4% for Model 2.

False negative rates ranged from 21.7% for Model 2 to 40% for Model 3. Results for our twelve-month analyses are included in Appendix A1. Similar results were observed for the twelve-month follow-up time point: Model 2 (ASQ + EHR) showed the highest sensitivity (0.746; Table A1) and AUC (0.843; Table A1).

**Table 2.** Binary Logistic Regression model comparison for accuracy of detecting a suicide related ED event three months after index event (*N* = 13,420)

| Model | Accuracy | Sensitivity | Specificity | Positive<br>Predictive<br>Value | False<br>Positive<br>Rate | False<br>Negative<br>Rate | AUC | Number of<br>features |
| --- | --- | --- | --- | --- | --- | --- | --- | --- |
| Model 1: ASQ Alone | 0.840 | 0.667 | 0.842 | 0.042 | 0.158 | 0.333 | 0.754 | 1 |
| Model 2: ASQ + EHR | 0.825 | 0.783 | 0.826 | 0.045 | 0.174 | 0.217 | 0.855 | 41 |
| Model 3: EHR alone | 0.832 | 0.601 | 0.835 | 0.036 | 0.165 | 0.399 | 0.775 | 40 |
| Model 4: EHR + ASQ-4 | 0.827 | 0.703 | 0.828 | 0.041 | 0.172 | 0.297 | 0.820 | 41 |

### Penalized Logistic Regression

**Table 3.** presents the comparison of model accuracy across the same four models but using penalized logistic regression to shrink the coefficients of the less contributive features. The results for Model 1 (i.e. ASQ screening alone) do not change because only one feature exists in the model. However, using the LASSO method, despite the substantially reduced number of features, we continue to see improvement in the sensitivity of the model when EHR is added to information from the ASQ. In fact, just adding one additional feature, a diagnosis of depression at index PED visit, results in this increased sensitivity: Model 2, the ASQ plus EHR has an accuracy of 0.799, sensitivity of 0.804 and AUC of 0.815; while Model 4, using only the 4^th^ question on the ASQ that reflects a self-report history of suicide attempted demonstrated higher accuracy (0.831), but lower sensitivity (0.659) and lower AUC (0.759). False positive rates ranged from 0.082 for Model 3 (i.e. EHR alone) to 0.201 for Model 2 (i.e. ASQ + EHR).

### Secondary Analysis

For the binary logistic regression, the inclusion of an index PED visit for suicide attempt or ideation coded in the EHR, improved the model accuracy of both Model 2 (ASQ + EHR; accuracy = 0.841) by 2% and Model 3 (EHR only; accuracy = 0.858) by 3% respectively. The AUC improved as well, with Model 2 increasing from 0.855 to 0.874 and Model 3 raising from 0.775 to 0.843 with the addition of this predictor. The largest improvement was observed for sensitivity for Model 3 (EHR only; elevating from 0.601 to 0.717). In summary, the secondary analysis depicted that if an EHR is capable of coding and utilizing this type of data, a model based only on EHR would be more sensitive, specific, and have a higher accuracy than the ASQ alone (see Appendix A2).

**Table 3.** LASSO model comparison for accuracy of detecting a suicide related ED event three months after index event (*N =* 13,420)

| Model | Accuracy | Sensitivity | Specificity | Positive<br>Predictive<br>Value | False<br>Positive<br>Rate | False<br>Negative<br>Rate | AUC | Number of<br>features |
| --- | --- | --- | --- | --- | --- | --- | --- | --- |
| Model 1: ASQ Alone | 0.840 | 0.667 | 0.842 | 0.042 | 0.158 | 0.333 | 0.754 | 1 |
| Model 2: ASQ + EHR | 0.799 | 0.804 | 0.799 | 0.040 | 0.201 | 0.196 | 0.815 | 2 |
| Model 3: EHR alone | 0.913 | 0.428 | 0.918 | 0.052 | 0.082 | 0.572 | 0.673 | 1 |
| Model 4: EHR + ASQ-4 | 0.831 | 0.659 | 0.833 | 0.039 | 0.167 | 0.341 | 0.759 | 2 |

## Discussion

### Main Findings

In this study we sought to compare a brief screening tool, the ASQ, to EHR-based predictive models for their accuracy at identifying individuals at risk of future suicide related ED visits. Our study found that the addition of EHR data to brief suicide screening in a PED setting considerably improves accuracy at identifying individuals at risk of future suicidal behaviors over the next three months. Our logistic regression findings indicate that relying solely on EHR data performs only slightly worse than implementing universal brief suicide screening. And while the best performing models were brief screening and EHR data, using just one item from a brief screener that asks about past history of suicide attempt (i.e., last question of the ASQ) and combining this data with EHR data, leads to substantial improvements in predictive accuracy compared to either screening or EHR models alone.

Given the potential to save lives by identifying those at risk of suicide in hospital settings, a substantial, but parallel effort has been made to develop brief screening measures^31,37^ and predictive models.^24–26,38^ To our knowledge, this is the first study to do a head to head comparison of these approaches. Our findings show that leveraging EHR data can be help improve predictive accuracy and utility when implementing universal screening procedures in a PED. Notably, model accuracy was only around 1% less for the EHR alone model (Model 3) compared to the ASQ (Model 1), though the latter remained 10% more sensitive. However, when added to the ASQ the EHR attributes (Model 2), improved sensitivity by 17.4% and AUC by 13.4% compared to the ASQ only model (Model 1).

Our secondary findings (Appendix 2) indicated that an index visit with diagnosis of suicidal behaviors was important in model accuracy. Inclusion of ideation or attempt at PED visits as a predictor improved model sensitivity by 9.8% and the AUC by 14% compared to the ASQ alone. Given known problems with documentation of ideation or attempt,^39^ our main findings indicate that simply asking one question from the ASQ in addition to the data gathered in a PED visit, is comparable to asking a full screening measure in terms of accuracy at identifying future suicide related PED visits. An advantage of screening, even with this one question, is it can give you the historical record and by-passes a potential limitation of EHRs – that if there is no documented history, the EHR by default considers this to mean there is no history, but we know that an absence of evidence is not evidence of absence.

Despite widescale recommendations to routinely screen for suicide risk among children and adolescents,^12,40^ instituting and implementing universal screening requires significant buy in, planning, and continued resources. First, instituting universal screening in a health system requires screening to be built into the work-flow of clinical care including protocols for addressing acute and nonacute screen positives.^41^ Given a lack of ready access to community mental health services, some hospital administrators can be reluctant to support institutionalizing this approach. Additional concerns involve the low positive predictive value of such screeners^20^ or operational modifications needed to implement these approaches. Implementation of screening requires training and ongoing oversight to ensure compliance.^18,42^ While our sample had high ASQ compliance rates, previous studies have shown lower compliance rates indicating a need for ongoing monitoring and quality improvement efforts.^43^ Notwithstanding, screening for suicide risk has shown promise at identifying patients at risk for suicide.^14,15^

Predictive modeling is considered potentially less burdensome from a human resource perspective. Instead of ongoing supervision to administer surveys, these modeling techniques leverage data analytics approaches that can be automated with ever advancing computing capacity through the EHR. However, suicide risk models may have considerable disadvantages. Some patients may feel that this approach is invasive of their privacy, particularly because they may or may not consent to having an algorithm applied to their data. Further, in-person screening may provide therapeutic benefits that would not be present with computational modeling, although the evidence for this is mixed.^44^

Both screening and predictive modeling approaches have limitations. The positive predictive value across all our models were quite low (e.g. 0.04-0.05). This is consistent with a recent systematic review by Belsher et al.^23^ focused on predictive models, findings from screening studies, and is reflective of the key challenge in suicide risk screening – suicide is a low base rate problem.^45^ Our false positive rates were considerably lower than those found in previous studies,^46^ but still represent potential inappropriate use of resources and/or a potential source of distrust or source of fatigue in procedures.^47^ While our work and the work of others show suicide risk models can accurately identify risk, depending on how they are operationalized, these models can generate additional workload for clinicians by necessitating response to false positives and strategies for responding to alerts on patients.^48^ Regardless of the approach, careful attention is needed to think about how a risk classification is communicated to the patient as both screening and predictive modeling may feel “out of the blue” to patients, particularly patients who present with medical complaints.

Despite these drawbacks, early and accurate identification of individuals at risk a critical component of a prevention framework. Implementation of either approach should be carefully studied. Developing optimal risk thresholds and care pathways is critical to realizing the value of these procedures in medical settings.^28^ Focusing on the net benefit of these approaches in a given setting may be one way to more clearly identify the optimal risk thresholds and support clinical decision making.^50^

Finally, few suicide risk prediction models have been developed in patient populations that represent racial and ethnic diversity. Our sample includes approximately two-thirds Black youth patients. Over the past decade, suicide rates are increasing at a faster rate among Black youth compared to any other racial or ethnic minority.^4^ This crisis needs to be addressed urgently; however, significant concerns exist about current approaches, including predictive models, which may exacerbate health disparities and/or potentially cause individual harm.^50^ Development and implementation of any risk identification strategy should not only consider net benefits, but also focus on incorporation of principles of distributive justice such as equalized outcomes, equal performance and equal allocation.^50^

### Limitations

The study has several important limitations worth noting. First, our outcomes did not include suicide deaths. What predicts ideation or attempts could be vastly different from what predicts future deaths by suicide. Moreover, our outcomes were not primarily defined by ICD codes, which may have resulted in some misclassification. Misclassification could have also contributed to too few cases identified given the potential overlap with drug overdoses. However, misclassification with ICD codes is a known issue in suicide research^39^ and a relative strength of this study is definition of outcomes through review of clinician notes. The generalizability of our findings may also be limited. First, data represents the patient population seen at the Johns Hopkins Hospital (JHH) PED. Generalizing these findings to rural populations, higher-income settings, and/or outpatient populations is not appropriate. Furthermore, our data is only representative of PED data captured at JHH, which excludes potential visits at other EDs. Finally, although we used rigorous methods for cross-validation, external validation of our models would further strengthen our findings.

### Conclusions

This study represents the first comparison of universal suicide screening and predictive modeling to identify individuals at risk of suicidal behavior in the next three and twelve months. We found that predictive modelling based on EHR may serve as a comparable approach when instituting a brief screening measure is not feasible. The combination of both brief screening, even in the form of a single item (e.g. ASQ4), and predictive modelling with EHR, substantially improves accuracy. Regardless of the method, careful attention should be paid towards optimizing the implementation of either approach, including focusing on net benefits and distributive justice principles.

## Data Availability

Data is not available publically.

## Appendix A1. Twelve-month prediction models

**A1.** Binary Logistic Regression model comparison for accuracy of detecting a suicide related ED event twelve months after index event (*N* = 10 205)

| Model | Accuracy | Sensitivity | Specificity | Positive<br>Predictive<br>Value | False<br>Positive<br>Rate | False<br>Negative<br>Rate | AUC |
| --- | --- | --- | --- | --- | --- | --- | --- |
| Model 1: ASQ Alone | 0.836 | 0.621 | 0.843 | 0.100 | 0.157 | 0.379 | 0.732 |
| Model 2: ASQ + EHR | 0.815 | 0.746 | 0.817 | 0.103 | 0.183 | 0.254 | 0.843 |
| Model 3: EHR alone | 0.818 | 0.621 | 0.823 | 0.090 | 0.177 | 0.379 | 0.789 |
| Model 4: EHR + ASQ-4 | 0.817 | 0.686 | 0.820 | 0.097 | 0.180 | 0.314 | 0.822 |

**Figure A1.**
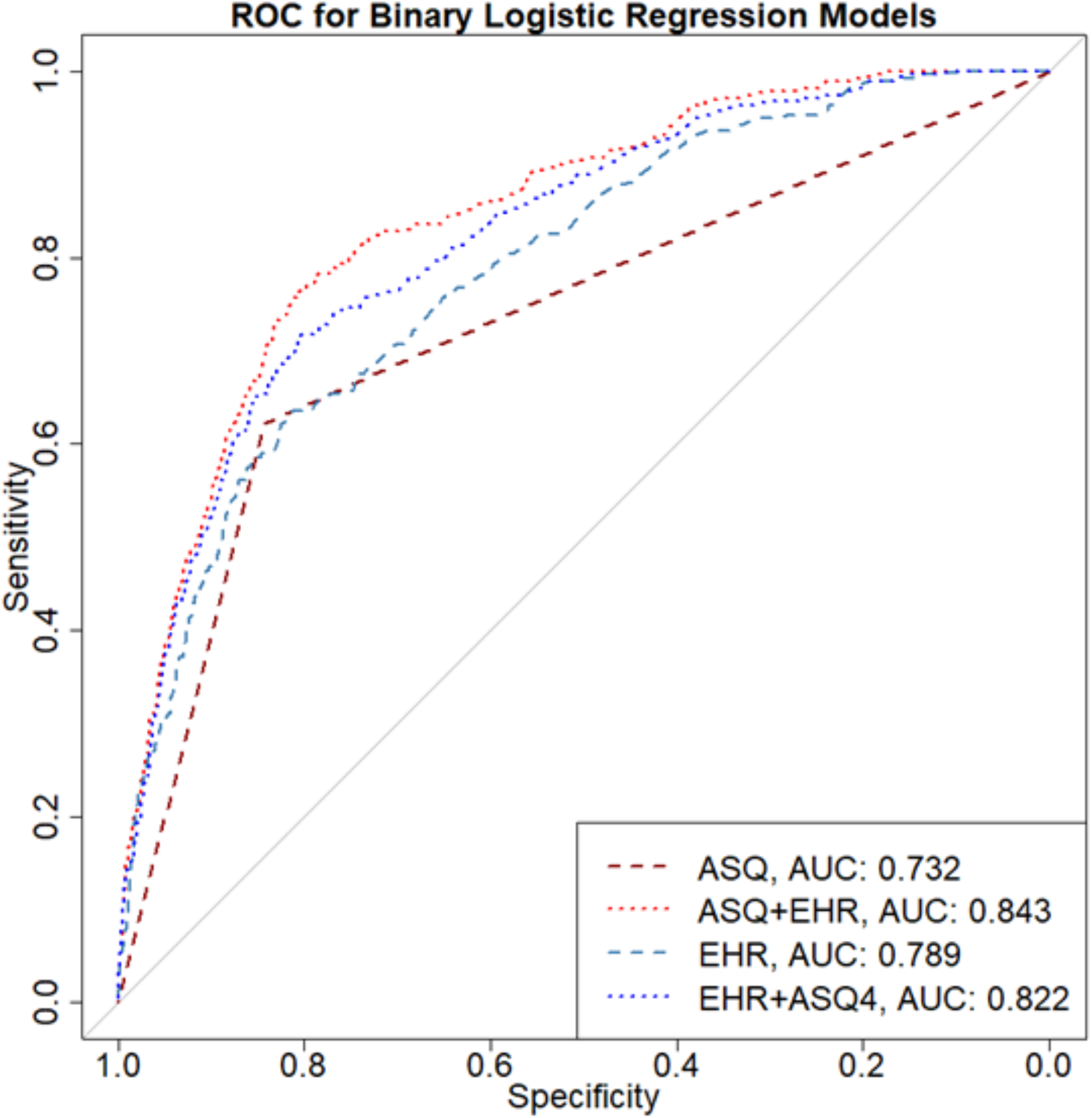
Receiver Operating Curve (ROC) comparing binary logistic regression models (*N =* 10 205) for prediction accuracy at 12 months

## Appendix A2. Model performance including suicide ideation or attempt at index PED visit as coded in the EHR

**Table A2.** Binary Logistic Regression model performance including the EHR marker for suicide ideation or attempt at index visit for detecting a suicide related ED event three months after index event (*N =* 13 420).

| Model | Accuracy | Sensitivity | Specificity | Positive<br>Predictive<br>Value | False<br>Positive<br>Rate | False<br>Negative<br>Rate | AUC |
| --- | --- | --- | --- | --- | --- | --- | --- |
| Model 1: ASQ Alone | 0.840 | 0.667 | 0.842 | 0.042 | 0.158 | 0.333 | 0.754 |
| Model 2: ASQ + EHR | 0.841 | 0.768 | 0.842 | 0.048 | 0.159 | 0.232 | 0.874 |
| Model 3: EHR alone | 0.858 | 0.717 | 0.860 | 0.051 | 0.140 | 0.283 | 0.843 |
| Model 4: EHR + ASQ-4 | 0.855 | 0.732 | 0.856 | 0.050 | 0.144 | 0.268 | 0.859 |

**Figure A2.**
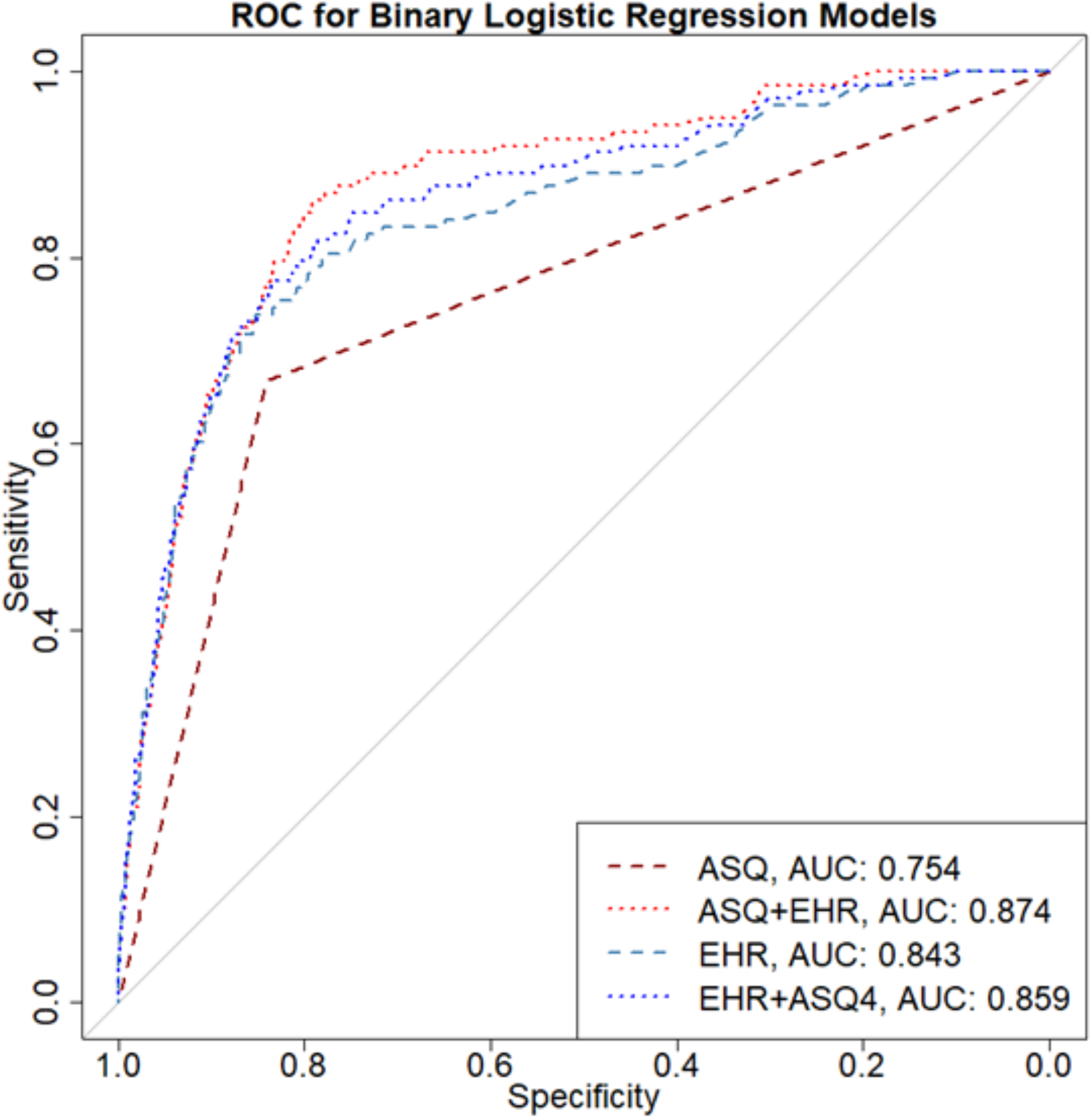
Receiver Operating Curve (ROC) comparing binary logistic regression models including the EHR marker for suicide ideation or attempt at index PED visit for prediction accuracy at 3 months (*N =* 13,420)

